# Estimation of COVID-19 case fatality ratio based on a bi-directional correction method

**DOI:** 10.1101/2020.05.02.20089144

**Authors:** Wanling Hu, Xiaoyun Liu, Tao Wang, Changlong Zhou, Dingfu Liu, Yuanming Zhang, Zhongli Hu, Ying Diao

## Abstract

The case fatality rate (CFR) can be used to predict the number of potential deaths in the epidemic and thus can reflect the appropriateness and quality of medical measures developed by public health. When a new disease breaks out, it is particularly important to accurately estimate the CFR. However, while the epidemic is still developing, the crude CFR is often lower than the true value and the hospital CFR is often higher than the true value due to differences in occurrence time, patient number, and treatment plans. Therefore, this study proposes a bi-directional correction method to estimate the CFR. COVID-19 data from China were used to evaluate this method. The results show that this method provides more accurate results than both the crude CFR and hospital CFR. Additionally, this method was used to estimate the CFR of COVID-19 in other countries, with an aim to provide a reference for prevention and control decisions for the COVID-19 epidemic and for the evaluation of medical efforts.

## 1. Introduction

At the end of December 2019, COVID-19, an epidemic disease caused by a novel coronavirus called SARS-CoV-2, broke out in Wuhan, China [1]. European and American countries generally failed to pay enough attention at the beginning of the outbreak. However, as the outbreak spread worldwide, the World Health Organization (WHO) characterized it as a pandemic on March 11, 2020. Similar to the SARS and MERS outbreaks, this serious epidemic disease was caused by a coronavirus. As of April 22, 2020, the number of people diagnosed with SARS-CoV-2 worldwide has exceeded 2.5 million, and the cumulative deaths have exceeded 170,000. Conversely, 8,098 were confirmed to have been infected by SARS worldwide, with a cumulative death toll of 774 and a case fatality rate (CFR) of 9.6%. As of January 2020, MERS had reported 2,519 confirmed infections worldwide and 866 deaths with a CFR of 34.3%. Obviously, the number of confirmed infections and deaths fromCOVID-19 is far higher than that of SARS and MERS. The published crude CFRs of COVID-19 are reported as 6.1% globally, with the highest CFR (8.3%) in Europe and the lowest (3.4-3.5%) in the Pacific and America. The CFR varies greatly in different countries, with the highest (14.3%) in France, CFRs above 12% in Italy and the United Kingdom, and 3.4% in the United States.

As the epidemic develops, there will be large differences in the calculated crude CFRs because of the inconsistent time that the epidemic has existed in various regions. For a certain region, the calculated crude CFR will display large deviations over time. That is, the CFR will be low at the beginning, but will gradually increase during the epidemic process, which may lead to incorrect judgments and decisions and may also affect the public’s confidence and trust in leaders [2]. As the severity of the COVID-19 epidemic has increased, and in the light of the strict government control measures, frequent medical resource problems in the news, and the increasing number of confirmed cases and deaths, people began to realize that COVID-19 is no longer an ordinary flu. Will COVID-19 exceed the CFR of SARS? Or even reach the CFR of MERS? At present, COVID-19 has been essentially curbed in China. Currently, more than 84,000 people have been diagnosed, nearly 93% have been cured and discharged, and more than 4,600 have died. This provides the basis and conditions for more objectively assessing the mortality of patients with COVID-19. Therefore, to improve the deviations in the estimates of the crude fatality rate caused by the continuously changing data during the epidemic, this study proposed a bi-directional correction method to estimate the CFR. This method was evaluated using the COVID-19 data from China. It further estimates the CFR of COVID-19 in other countries. The research results not only provide a reference for the evaluation of medical treatment and decision-making for epidemic prevention and control, but they also have important significance for stabilizing society and people’s feelings regarding the epidemic.

## 2. Methods and data sources

### 2.1 Definition, model, and correction of CFR

The CFR is defined as the number of people or animals who die from a disease during a certain period of time out of the total number of patients or animals infected. For human diseases, “death” is the end of medical treatment due to illness, and “cure” is defined as discharge from the hospital after the end of treatment. Consider the following scenario: if the epidemic situation of a certain disease has ended and all patients have finished treatment, suppose the cumulative number of people cured is ∑x_i_ and the cumulative number of deaths 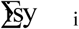. Then, the cumulative number of confirmed cases is (∑T_i_) = (∑x_i_ + ∑y_i_), as follows:

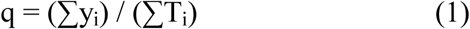

or,

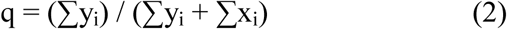

Here, i = 1, 2, n, but the epidemic has ended. After the epidemic is over, the crude method can be a good estimate of the CFR. Additionally, formulas (1) and (2) give exactly the same value.

However, while the epidemic is still occurring, calculating the CFR involves many complicated factors. The WHO used two methods to estimate the SARS CFR using accumulative case data [3]. One is the widely adopted estimator of the CFR, commonly known as the crude CFR, which underestimates the true CFR as the epidemic continues. The crude CFR is calculated by formula (1) using the cumulative number of deaths 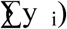) on day (n) and the cumulative number of diagnosed cases (∑T_i_) on the same day (n), as follows:

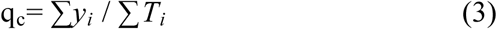

Here, i = 1, 2, …, n.

The second method involves using only the final number of deaths and the number of discharges at the time of the epidemic, that is, it substitutes the cumulative number of discharges (∑x_i_) and cumulative deaths (∑y_i_) of day (n) into formula (2). This results in an overestimation of the CFR during the ongoing epidemic. For convenience, this method is called the hospital CFR, and it is calculated as:

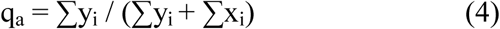

Here, i = 1, 2, …, n.

In general, the natural procedures for diseases such as SARS and COVID-19follow the general pattern of viral infection-onset-treatment/hospitalization-diagnosis-death or rescue-discharge, but different diseases have different courses [4]. To utilize easy-to-obtain statistical data, special consideration is given to the “confirmed-death or rescue-discharged” time period, which involves only three time points: confirmed, death or rescue, and discharged (Figure 1). For example, the period from diagnosis of SARS cases to death is approximately 10 days [5], which indicates that the death cases observed until a certain day are generally patients who were diagnosed 10 days ago, and the cumulative number of cases includes the number of new cases in the last 10 days. In fact, most of these patients have not reached the time point of “death or rescue” caused by the disease. In formula (3), this part of the population should not be included in the denominator of the CFR calculation. The same is true for COVID-19 case data. Considering its generality, that is, because is a time difference (k days) between when the patients confirmed and when they died (Figure 1), formula (3) is corrected as:

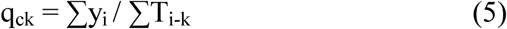

**Figure 1.**
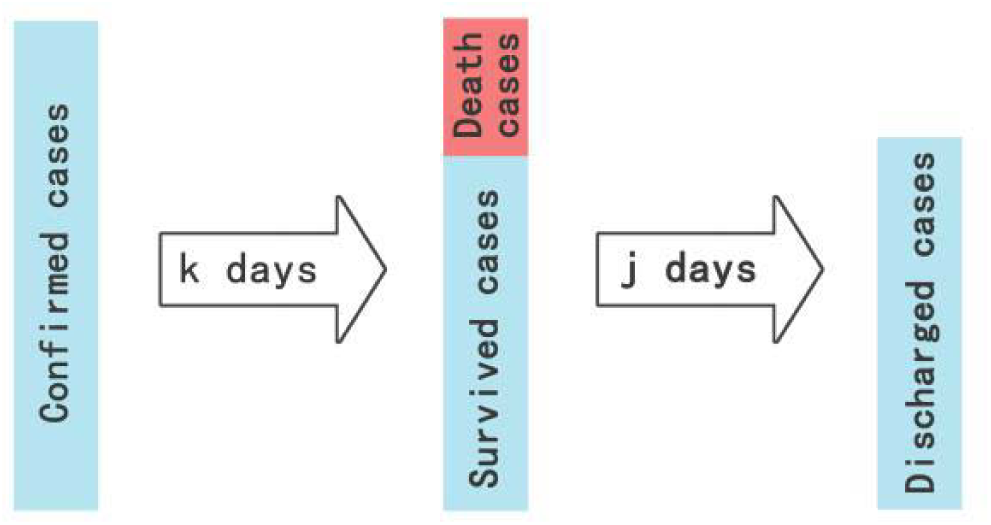
A course schematic diagram of COVID-19

Here, i = 1, 2, …, n.

When evaluating this from the perspective ofCOVID-19, according to clinical research, the early symptoms are mild. After a week, the disease may worsen or the patient may even die; however, as long as the patient passes the most dangerous period of time, he can gradually recover. Moreover, in China, according to the treatment plan, cured patients must wait for all symptoms to disappear and then test negative forSARS-CoV-2 nucleic acid twice before classified as cured and being discharged. In other words, the patients with COVID-19 who died and those who were discharged on a particular day were not admitted to the hospital at the same time. For patients who were confirmed at the same time, there was a time difference (j days) between when those patients were either discharged or died. The same was true for SARS case data. Without a loss of generality, to correct formula (4), we propose an estimator of CFR:

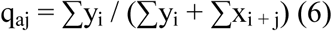

Here, i = 1, 2, …, n.

Obviously, formulas (3) and (4) are special cases of formulas (5) and (6) when k = 0 and j = 0, respectively.

### 2.2 Estimated CFR with the bi-directional correction

According to the structure of formulas (5) and (6), the estimated values of q_ck_ and q_aj_ calculated using formulas (5) and (6), respectively, can be combined with the cumulative death toll of a certain day and observed in the “Golden Cross”, that is, the interval where q_ck_ = q_aj_ appears should contain the true CFR. It is clear that the three cumulative data points corresponding to the three time points involved in the “diagnosis-death or rescue-discharge” periods have the following logical relationship:

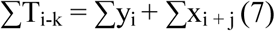

It is easy to derive q_ck_ = q_aj_ from equation (7).

Since k and j are both variable, some conditions must be set for estimating the true mortality rate. These conditions are easy to understand and are in line with statistical principles and clinical laws, for example: q_ck_ < q_a0_, q_aj_> q_c0_; k < j; etc. Therefore, we propose the following principles and steps of the bi-directional correction method to estimate the mortality rate:

1. Take k = 0, 1, 2, …, 15 and j = 0, 1, 2, …, 20, and enter them into formulas (5) and (6), respectively, calculate the values of q_ck_ and q_aj_, according to q_ck_ < q_a0_ and q_aj_> q_c0_, and determine the maximum values of k and j (k_max_ and j_max_, respectively), which constitute the interval of q_ck_ = q_aj_.

2. According to the q_ck_ = q_aj_ interval composed of k_max_ and j_max_, calculate the mortality using the values of the mortality (time series) corresponding to k = 0, 1, 2, …, k_max_ and j = 0, 1, 2, …, j_max_. Find the Manhattan distance or Euclidean distance between the sequences, and determine the two fatality sequences with the smallest distance (they are derived from q_ck_ and q_aj_, respectively).

3. Average the two fatality series with the smallest distance to obtain an estimated value series. Two types of averages can be considered: 1) arithmetic average and 2) weighted average, “weight” is taken as k / (j + k) and j / (j + k), respectively, in which k and j correspond to the two CFR sequence with the smallest distance.

For convenience, we can call the method of determining k and j values as the real-time CFR sequence distance minimum determination method”.

### 2.3 Data source and description

China data sources: (1) The data for mainland China (except for Hong Kong, Macao, and Taiwan) were derived from the data released by the Hubei Provincial Health Commission and the National Health Commission from January 20 to April 22, 2020, including cumulative and new diagnoses, deaths, and discharges. (2) However, the Wuhan Municipal Government revised the data on April 17 because the revised data (an increase of 1290 cases) in the cumulative death toll was not included in the previously published cumulative number of confirmed cases, so it was not included in the calculation in this study (TableS1).

Foreign data sources: (1) The cumulative number of diagnoses and the number of deaths come from the data published by the WHO and span from the date of the announcement of the cumulative deaths (February 3, 2020) to April 9. (2) The cumulative number of cured people in eight countries comes from the Tencent News Network. (3) Because Tencent News Online does not have data on the cumulative number of cured people in “Global except China”, data from Phoenix News Online were used. The cumulative number of cured people has not been updated, but the corresponding Phoenix News online data were updated for the same time period; thus, the data from Phoenix News Online were used (TableS2).

ChinaCOVID-19 data were used to evaluate and verify the bi-directional correction method for CFR proposed in this paper, and this method was also used to estimate the global CFR and the CFR in eight countries with the most severe epidemics.

## 3 Results

### 3.1 Estimation and comparison of the COVID-19 CFR in China

#### 3.1.1 Bi-directional correction estimates

According to geographical divisions, China’s COVID-19 case data can be divided into three groups: Wuhan, Hubei except Wuhan, and national except Hubei. The data do not overlap with each other. In all regions, the characteristics of the cases are essentially the same, and the medical treatment methods are essentially the same, so the course of the disease is similar. The three sets of data and the national data were analyzed according to the bi-directional correction method to estimate the CFR. The intermediate results are shown in Table 1. Based on these results, the average of the mortality rate series corresponding to k and j in Table 1 was calculated. The time series of the estimated value of the CFR of the COVID-19 in China is shown in Figure 2, in which the relationship between the time series of various CFR estimates clearly displays. The CFR (arithmetic mean) estimated by the bi-directional correction is essentially in the middle, which is slightly higher than the crude CFR. The CFR of the bi-directional correction method makes up the deficiencies of the lower estimates for the crude CFR so as to produce a more accurate estimate. Of course, all the estimates tend to reach the same value in the end, that is, the “final CFR”.

**Table 1.** The k and j values of China COVID-19 data

|  | k/j | Wuhan | Hubei except Wuhan | China except Hubei | China |
| --- | --- | --- | --- | --- | --- |
| | $k_{\max}$ | 8 | 4 | 7 | 8 |
| | $j_{\max}$ | 11 | 11 | 11 | 11 |
| Manhattan Distance | k | 3 | 4 | 6 | 3 |
|  | j | 11 | 11 | 11 | 11 |
| Euclidean Distance | K | 3 | 4 | 3 | 3 |
|  | j | 11 | 11 | 11 | 10 |

**Figure 2.**
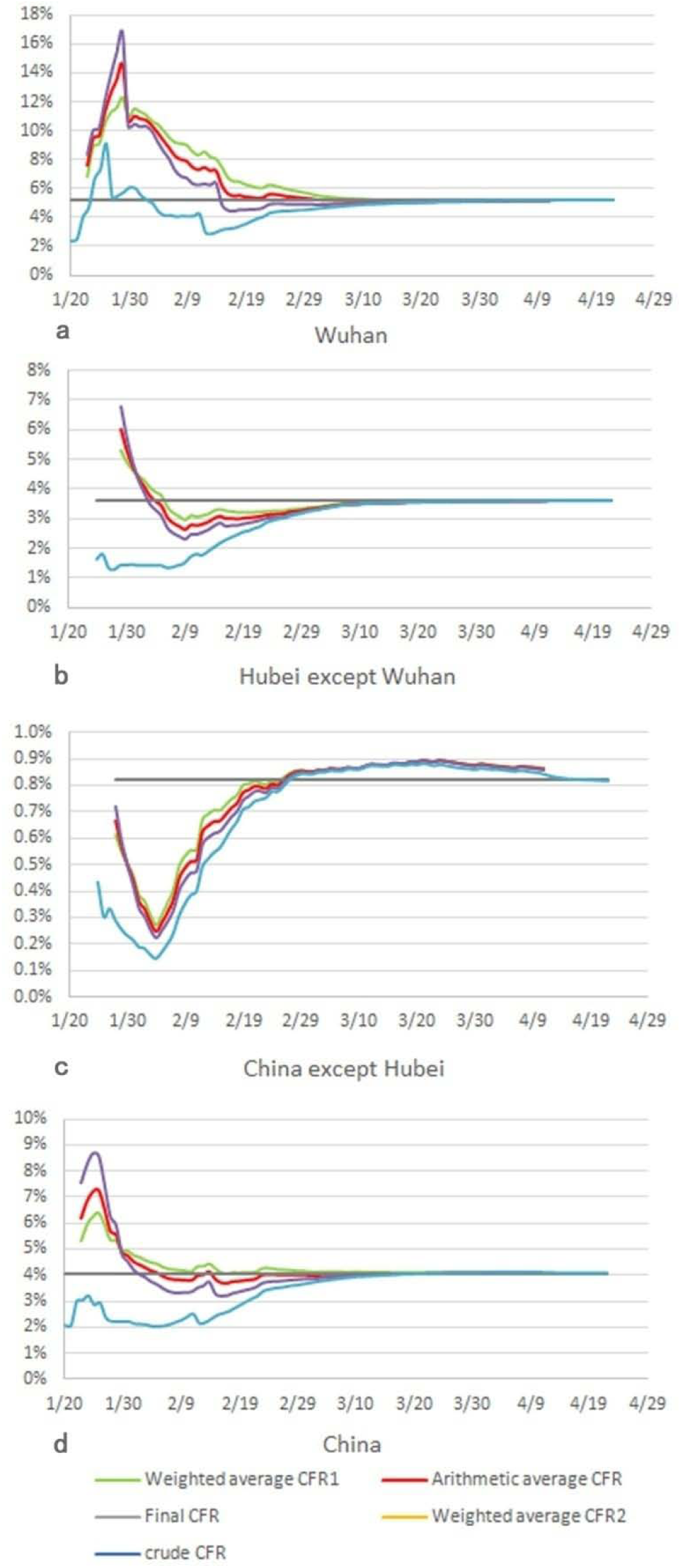
Time series of CFR estimates for COVID-19 in China

#### 3.1.2 Comparison of the estimated CFR with the final CFR

To compare the differences in the arithmetic mean CFR of the bi-directional correction method, the weighted average CFR, the crude CFR, and the final CFR (q_z_), two evaluation indicators were used: (1) the Manhattan distance, i.e., absolute difference = ∑ | q-q_z_ | / n; (2) relative difference = ∑ (q -q_z_) / n. In addition, the weighted average CFR from the bi-directional correction is divided into two conditions: the calculated CFR is recorded as the weighted average CFR 1 using the weight of q_ck_ is k / (j + k) and the weight of q_aj_ is j / (j + k); conversely, the calculated CFR recorded as the weighted average CFR 2 using the weight of q_ck_ is j / (j + k)and the weight of q_aj_ is k / (j + k). The calculation results are listed in Table 2.

**Table 2.** Comparison of CFR estimates and final CFR of COVID-19 in China

| Area | Final CFR<br>($q_z$) | Evaluation index | Crude<br>CFR | Arithmetic mean<br>CFR | Weighted average<br>CFR1 | Weighted average<br>CFR2 |
| --- | --- | --- | --- | --- | --- | --- |
| Wuhan | 5.16% | Relative<br>difference | -0.0047 | 0.0133 | 0.0157 | 0.0108 |
|  |  | Absolute<br>difference | 0.0072 | 0.0136 | 0.0159 | 0.0136 |
| Hubei except | 3.61% | Relative | -0.0077 | -0.0015 | -0.0008 | -0.0021 |
| Wuhan |  | difference |  |  |  |  |
|  |  | Absolute | 0.0077 | 0.0033 | 0.0025 | 0.0041 |
|  |  | difference |  |  |  |  |
| China except | 0.82% | Relative | -0.0013 | -0.0006 | -0.0005 | -0.0007 |
| Hubei |  | difference |  |  |  |  |
|  |  | Absolute | 0.0018 | 0.0012 | 0.0011 | 0.0014 |
|  |  | difference |  |  |  |  |
| China | 4.05% | Relative | -0.0075 | 0.0021 | 0.0026 | 0.0012 |
|  |  | difference |  |  |  |  |
|  |  | Absolute | 0.0076 | 0.0031 | 0.0026 | 0.0053 |
|  |  | difference |  |  |  |  |

Note: The "final CFR ( $q_z$ )" listed in the table is also an estimate. Based on April 9, the number of deaths in before 10 days of this date is $d$ , the number of patients cured is $c$ , and the number of patients currently in hospital is $y$ , then the number of patients who may die in hospital at the end is $y * d / (c + d)$ ; and this number of deaths plus the current cumulative number of deaths, divided by the cumulative number of confirmed cases, is called the final CFR ( $q_z$ ).

Overall, for the Wuhan data, the difference between the crude CFR and the final CFR was the smallest; conversely, for the Hubei except Wuhan, China except Hubei, and China data, the difference between the crude CFR and the final CFR was the largest, which indicates that the CFR estimated by the bi-directional correction is closer to the true CFR in most cases. Figure 3 shows the differences and trends of the weighted average CFR 1, the weighted average CFR 2, the arithmetic mean CFR, the crude CFR, and the final CFR of COVID-19 in China. In the early stage of the epidemic in Wuhan (Fig.3 a), when several estimates deviated from one another, the crude CFR appeared closer to the final CFR; but 20 days after the epidemic had begun, the bi-directional correction estimate was closer to the true CFR than the crude CFR.

**Figure 3.**
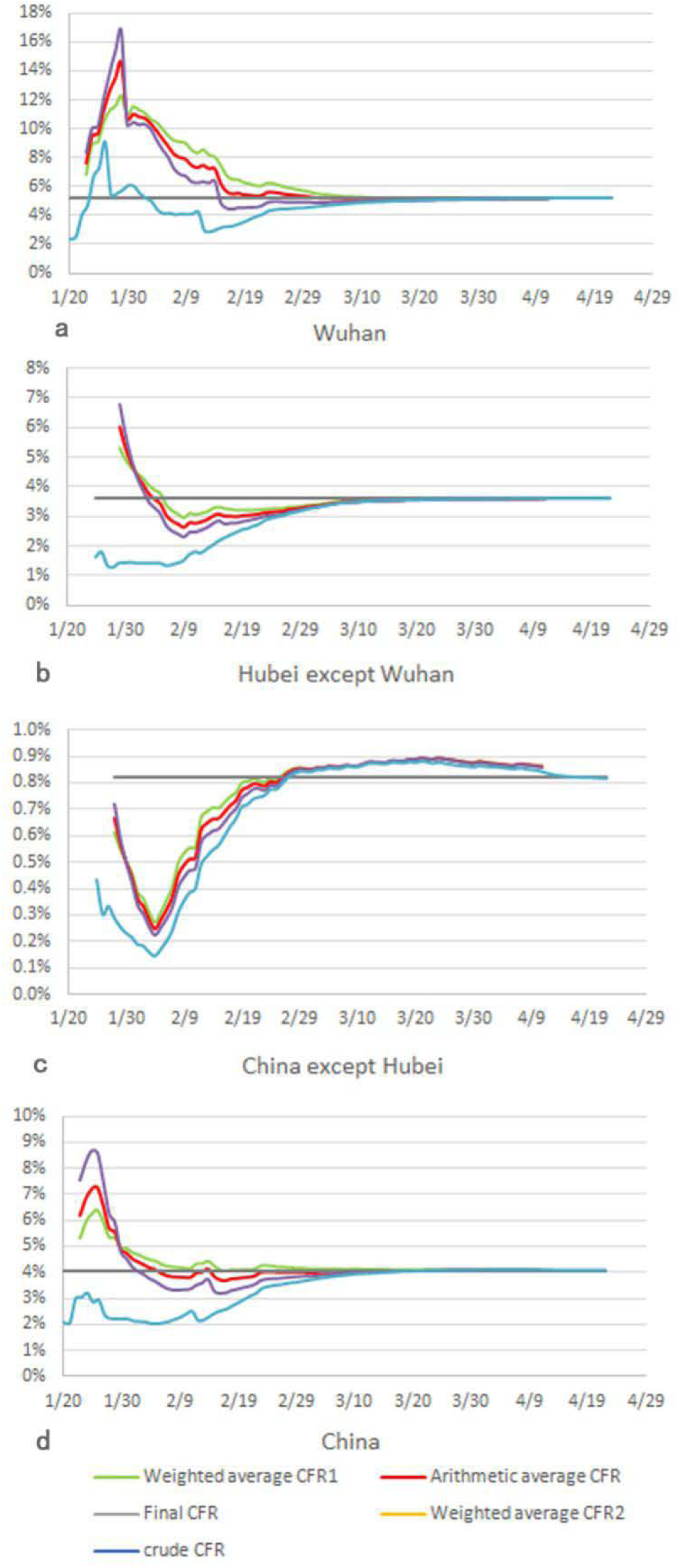
Trends of the CFR estimates calculated by four methods for COVID-19 in China

#### 3.1.3 The relationship between the statistical distribution of the new daily data of COVID-19 in China and the k and j values

Using the daily data of newly diagnosed, discharged, and dead patients in China, the trend of newly diagnosed, discharged, and dead cases of COVID-19 in China from January 20 to April 9 can be evaluated (Figure 4).

**Figure 4.**
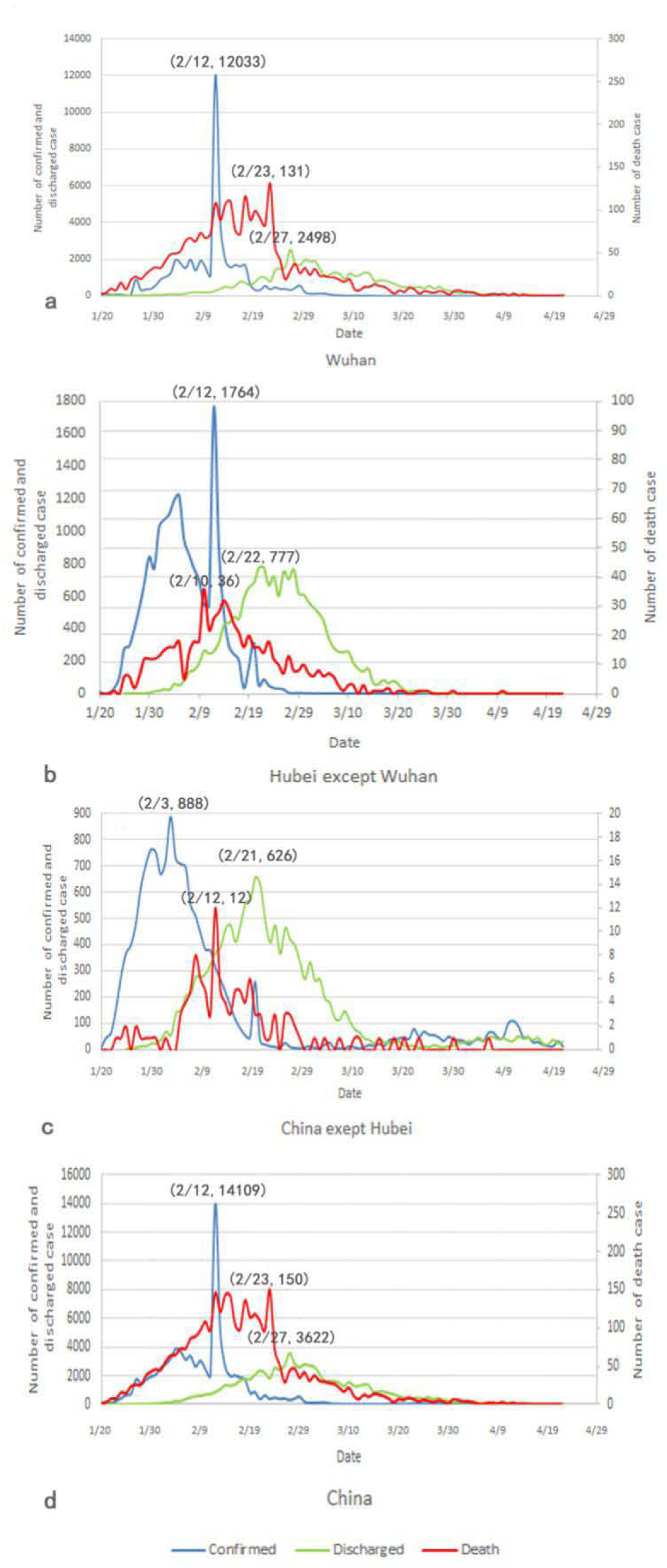
Trends of theconfirmed, discharged and dead Cases of COVID-19 in China

In Figure 4, the first peak in data occurs for the number of diagnosed cases, followed by peaks for death and discharge. This is a good verification of the logical relationship given by formula (7). First, the peaks appeared earlier in areas with mild epidemic and the peaks appeared later in areas with heavy epidemics. Second, for the interval time from the diagnosis peak to the death peak was6, 9, 9, and 6 days in Wuhan, Hubei except Wuhan, China except Hubei, and China data, respectively; and the interval time from the death peak to the discharge peak was10, 8, 8, and 10 days, in Wuhan, Hubei except Wuhan, China except Hubei, and China data, respectively. Therefore, the time from diagnosis to discharge may be approximately17 days.

Here, another statistical method can be used to determine the k and j values, which is tentatively called “the method for determining the k and j values by the peak interval time”. However, this method may only be applied in the middle and later periods, even after various peaks of the epidemic situation data appear. Of course, this statistical distribution result given by China’sCOVID-19 data is definitely of reference value for verifying the theoretical method of estimating the CFR and analyzing foreign COVID-19 data. Compared with the results in Table 1, the peak interval time given here corresponds to a larger k value and a smaller j value. If the CFR is estimated based on this method, the estimated CFR is higher.

### 3.2 Estimated global mortality rate of COVID-19

According to the bi-directional correction method and procedure for estimating the CFR, the statistical analysis of the data from eight countries around the world is shown in Table 3. Since the k and j values calculated by foreign data are too diverse, the additional filter k≥3 must be added. For comparison, the table also lists the current crude CFR. Because there is a time lag of j days, the estimated mortality rate of the bi-directional correction is actually the mortality rate of j days ago, so Table 4gives the estimated results for March 20 and March 30, respectively, while the estimation result for April 9 is only given by formula (5) with a one-directional correction, that makes a slightly worse accuracy for the estimate value than bi-directional correction, but more accurate than the crude CFR. In addition, the trend of the CFR in the eight countries is shown in Figure 5 (TableS3).

**Table 3.** Comparison of COVID-19 CFR in several countries

| Country | Time lag |  | March 23 |  |  | April 2nd |  |  | April 12 |  |  | April 22 |  |  |
| --- | --- | --- | --- | --- | --- | --- | --- | --- | --- | --- | --- | --- | --- | --- |
|  | k | j | Crude CFR | Arithmetic mean CFR | Withdrawal rate | Crude CFR | Arithmetic mean CFR | Withdrawal rate | Crude CFR | Arithmetic mean CFR | Withdrawal rate | Crude CFR | Arithmetic mean CFR | Withdrawal rate |
| Italy | 5 | 7 | 9.26% | 23.49% | 21.14% | 11.90% | 24.20% | 27.13% | 12.79% | 22.46% | 34.15% | 13.40% | 14.59% | 41.45% |
| Spain | 4 | 7 | 6.02% | 12.41% | 15.03% | 8.86% | 14.20% | 31.04% | 10.10% | 14.80% | 46.62% | 10.42% | 11.32% | 50.84% |
| Iran | 3 | 7 | 7.79% | 10.56% | 44.36% | 6.38% | 9.01% | 38.89% | 6.22% | 6.98% | 66.12% | 6.25% | 6.55% | 78.14% |
| South Korea | 4 | 14 | 1.17% | 1.48% | 33.87% | 1.69% | 1.95% | 60.11% | 2.04% | — | 72.13% | 2.23% | 2.23% | 79.62% |
| Japan | 6 | 15 | 3.68% | 5.85% | 28.68% | 2.39% | 4.75% | 22.19% | 1.45% | — | 12.74% | 2.41% | 2.30% | 14.20% |
| Germany | 4 | 9 | 0.38% | 0.92% | 2.08% | 1.19% | 1.65% | 26.84% | 2.22% | 2.71% | 46.97% | 3.35% | 3.55% | 68.69% |

| y |  | % |  |  |  |  |  |  |  |  |  |  |  |  |
| --- | --- | --- | --- | --- | --- | --- | --- | --- | --- | --- | --- | --- | --- | --- |
| USA | 3 | 1<br>2 | 1.27<br>% | 3.99% | 1.84% | 2.05% | 5.76% | 5.87% | 3.76% | —— | 9.68% | 4.84% | 5.41% | 14.30% |
| France | 4 | 1<br>0 | 4.26<br>% | 6.60% | 18.17% | 7.14% | 11.96% | 26.79% | 14.89<br>% | 21.81% | 43.64% | 17.88<br>% | 19.20% | 52.18% |
| Global<br>except<br>China | 4 | 1<br>0 | 4.47<br>% | 8.58% | 15.50% | 5.19% | 9.52% | 20.20% | 6.36% | 10.94% | 26.70% | 6.89% | 7.92% | 33.75% |

**Figure 5.**
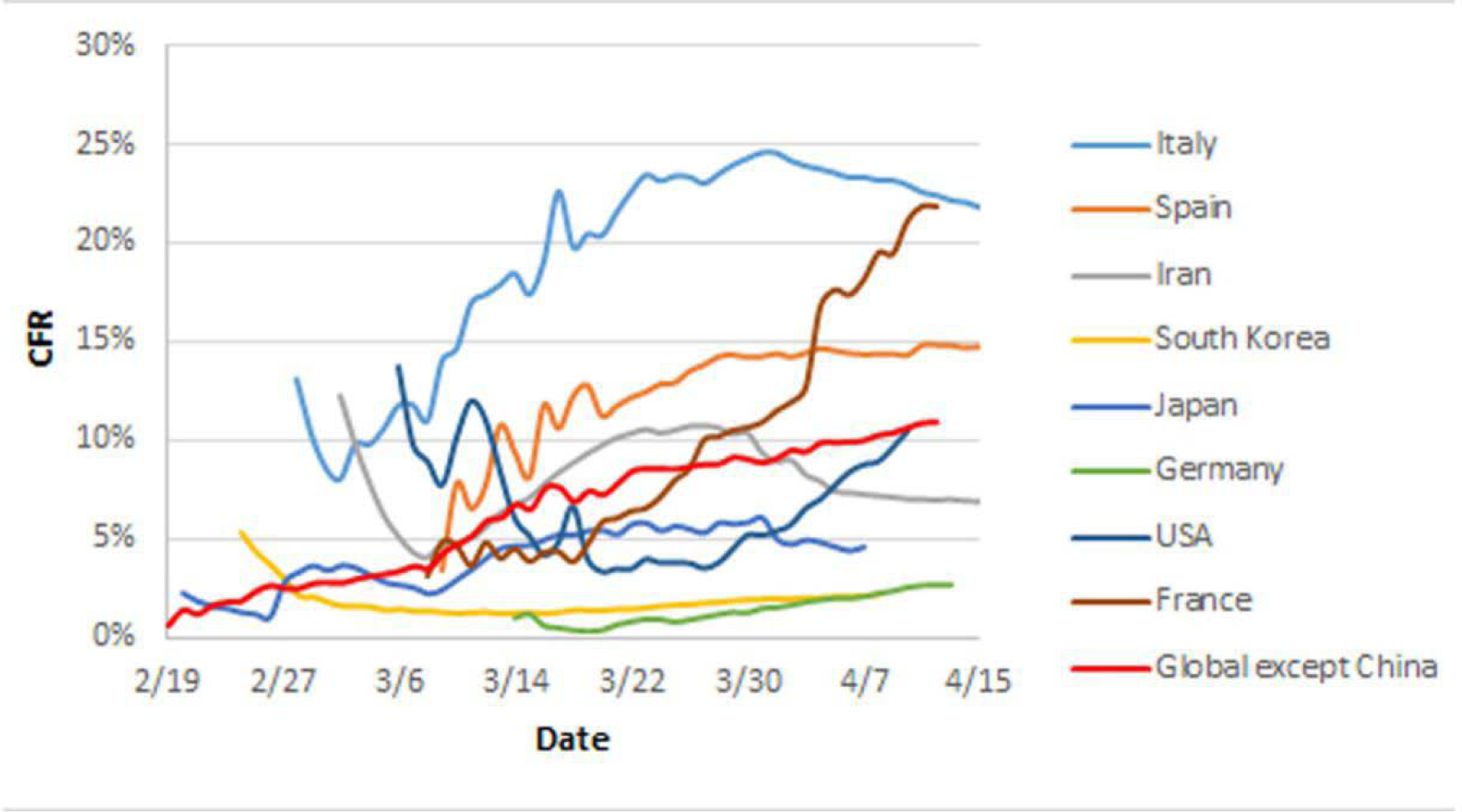
Trends of the CFR estimatesof COVID-19 worldwide

As shown in Table 3, the time lags of K are relatively similar and range from three to five days in the eight countries, but the time lag of j is higher and ranges from 7 to 15 days. Since j depends on the cure data, the difference of j is related to the number of discharges/recoveries announced by each country and the timeliness of the announcement. The disease termination rate or case withdrawal rate reflects the overall progress of epidemic treatment and the epidemic situation. For countries with a disease termination rate of close to or greater than 50%, such as South Korea and Iran, the crude CFR is close to our estimates, which also shows that these two estimates are approaching the final CFR. For countries with a low disease termination rate and where the epidemic situation is more serious, the CFR estimated by the bi-directional correlation method has risen and fallen in time to reflect the epidemic situation at that time. For example, the estimated CFR of Italy was lower on April 9 than that reported on March 20, indicating that the epidemic is improving. The crude CFR cannot show this trend because the crude CFR increases from low and high over time, which is one of its characteristics. From the perspective of change overtime, the change in our estimates is smaller than the change in the crude CFR. For example, our estimate of the CFR in Germany has a maximum difference of approximately seven-fold, and the difference in the crude CFR is more than ten-fold; moreover, our estimate of the worldwide CFR except for China has been relatively stable and has only increased by 0.6%whereasthe crude CFR has increased by approximately2%. The increase in CFR estimates in most countries indicates that the epidemic continues to worsen, and the range of the increase indicates the intensity of the outbreak.

## 4 Discussion

### 4.1 The estimated CFR of COVID-19 in China using the bi-directional correction method

TheCOVID-19 epidemic is essentially over in China, and the CFR has almost reached its final value. By using this complete epidemic data, we can evaluate or compare different CFR estimation methods. The CFR estimated by the bi-directional correction method is closer to the final CFR than the crude CFR. In the early stages of the Wuhan epidemic (approximately the first 20 days of the epidemic), those estimates deviated largely from one another. The crude CFR appeared closer to the final CFR, but in fact, the estimated CFR of the bi-directional correction may be closer to the true CFR than the crude CFR. This occurs because for an unknown novel epidemic, with the occurrence and development of the epidemic, the true CFR should not be a horizontal line, nor a curve from bottom to top (low to high); instead, the dynamic CFR curve should be an “Ω” curve with steep fronts and gradual backs [6]. We think this may be more consistent with actual clinical conditions, that is, with the characteristics of a steep front and gradual back. Therefore, the estimates of the bi-directional correction are closer to such a curve, which unfortunately cannot be verified because it is almost impossible to give the exact number of infections, deaths and so on at each time point of the epidemic to calculate true CFR.

In theory, the bi-directional correction method takes the average value of the corrected confirmed CFR q_ck_ and the corrected in-patient CFR q_aj_ to estimate the CFR. q_ck_ is known to be underestimated and q_aj_ is overestimated. On average, the two exactly hedge the deviations in their independent estimates. Therefore, the estimated CFR of the bi-directional correction method proposed in this article may be able to predict the severity of the outbreak early for confirmed hospitalizations. Of course, in situations when less than 100% of confirmed cases are hospitalized, the hospitalization rate must be considered when calculating the average values of q_ck_ and q_aj_ to determine the estimated CFR.

### 4.2 The time delay (k, j) in the bi-directional correction estimation method

When estimating the CFR using accumulative case data, if the time lag (k, j) is taken into account for correction calculations, statistical methods or clinical experience, or a combination of both, may be used to determine the k and j values. Based on clinical observations, Lei (2003) believed that the time from diagnosis to death in SARS cases was approximately 10 days (5). Thus, the determination of k = 10 was based on clinical experience. According to statistical theory, Hu et al. (2020) used “the minimum coefficient of variation of the daily estimated CFR” as the selection metrics, that is, they used the real-time CFR minimum coefficient of variation to determine the best values of j, which were 8, 10, 10, and 12 in Wuhan, Hubei except Wuhan, China except Hubei, and China, respectively [7]. According to the bi-directional correction method and program proposed in this paper, the analysis of the method and procedure is based on the real-time CFR sequence distance minimum determination method, in which k = 3 or 4 and j = 11 were determined, which is essentially consistent with actual clinical conditions. China adopts unified medical treatment standards, particularly the principles that all suspected and confirmed patients should be admitted to the hospital and all confirmed patients should be treated, that is, once the diagnosis is confirmed and regardless of whether it is mild, the patient must be isolated at a designated medical place. Thus, because all confirmed patients had complete records of hospitalization to recovery and discharge, these data can yield more consistent k and j values.

Different countries have different attitudes, prevention and control strategies, and medical treatment methods for COVID-19, and the nature or quality of published statistical data also differ; these differences will affect data analysis and results and cause deviations. For example, due to the different diagnosis and treatment standards abroad, most countries with mild patients require home isolation. After asymptomatic isolation for several days, they can be regarded as cured, without the need for viral nucleic acid testing again; thus, the course data of these COVID-19 patients are incomplete. In some countries, the case data are incomplete or only record the entry of patients and ignore the exit; thus, the withdrawal rate is very low, or the number of confirmed cases does not equal the sum of the number of deaths, cures, and hospital treatment. Therefore, the estimated k value and j value are diversified.

Further research is required to determine whether or how the third method mentioned in Section 2.1.3 of this paper, that is, the method for determining the k and j values by the peak interval time, or referring to the information given by the cumulative data curve of the cases, can be used to estimate the actual CFR.

### 4.3 The CFR of COVID-19 in foreign countries

The crude CFR was low at the beginning of the epidemic, which may have created serious misjudgment by decision-makers. For example, the United Kingdom first proposed the strategy of herd immunity; the US President also believes that the COVI-19 CFR will not exceed 1%, which is lower than the ordinary flu. Unfortunately, these original judgments were wrong, and those leaders missed the effective time for epidemic containment. Our estimate was relatively high from the beginning and may be closer to the true CFR. This information may remind policy makers to be vigilant and take appropriate epidemic prevention measures to reduce the impact of the epidemic on society.

From the perspective of the valuation curve, in the early stage of the epidemic, most countries were in line with China’s trend, and there was a large fluctuation in the CFR. This may be related to inadequate medical resources during the early stages and the intensity of the outbreak. With the use of strict prevention and control measures, the estimation of the CFR will stabilize. It is worth noting that regardless of the crude CFR and our estimates, the current epidemic situation in European countries may be becoming serious. The existing medical methods and prevention and control measures have not been able to adequately stop the epidemic.

The latest research shows that SARS-CoV-2 has been differentiated into three types labeled A, B, and C, of which type C is the predominant type in Europe [8]. At present, the rapid increase in the number of confirmed cases in Europe and the high CFR, in addition to failing to effectively block the spread of the virus, may be related to the change in the pathogenicity caused by the differentiation of the virus.

## Data Availability

The availability of all data in the manuscript are list in the tables and supplementary tables.

## Supplementary Materials

Table S1: Case number of COVID-19 in China

Table S2: Case number of COVID-19 in some countries

Table S3: The CFR estimated by bi-directional correction method for some countries

## Author Contributions

Y.D. conceived the study, W.H., X.L. and T.W. analyzed using models. C.Z., Y.Z. and D.L. collected the data. All authors participated in the study design. Y.D. and Z.H. drafted the manuscript. All authors gave comments on the earlier versions of the manuscript. All authors edited the manuscript and approved the final version.

